# Passively Sensing SARS-CoV-2 RNA in Public Transit Buses

**DOI:** 10.1101/2021.06.02.21258184

**Authors:** Jason Hoffman, Matthew Hirano, Nuttada Panpradist, Joseph Breda, Parker Ruth, Yuanyi Xu, Jonathan Lester, Bichlien Nguyen, Luis Ceze, Shwetak N. Patel

## Abstract

Affordably tracking the transmission of respiratory infectious diseases in urban transport infrastructures can inform individuals about potential exposure to diseases and guide public policymakers to prepare timely responses based on geographical transmission in different areas in the city. Towards that end, we designed and tested a method to detect SARS-CoV-2 RNA in the air filters of public buses, revealing that air filters could be used as passive fabric sensors for the detection of viral presence. We placed and retrieved filters in the existing HVAC systems of public buses to test for the presence of trapped SARS-CoV-2 RNA using phenol-chloroform extraction and RT-qPCR. SARS-CoV-2 RNA was detected in 14% (5/37) of public bus filters tested in Seattle, Washington, from August 2020 to March 2021. These results indicate that this sensing system is feasible and that, if scaled, this method could provide a unique lens into the geographically relevant transmission of SARS-CoV-2 through public transit rider vectors, pooling samples of riders over time in a passive manner without installing any additional systems on transit vehicles.

**Synopsis:** Passive sensing of viral presence on urban transit infrastructure is proven, with forward-looking benefits for tracking pandemic spread.

## Introduction

The global pandemic of COVID-19 has exceeded 33 million reported cases in the US and 168 million confirmed cases worldwide as of May 28th, 2021. ^1,2^ The virus causing COVID-19, SARS-CoV-2, is primarily transmitted through airborne respiratory droplets via face-to-face contact^3–5^ with asymptomatic or pre-symptomatic infected individuals.^6–11^ Therefore, disease monitoring via viral presence testing is essential for managing potential outbreaks. Current disease monitoring is focused primarily on testing individual members of the population. However, frequent widespread testing across the entire population can be cost-prohibitive in many communities, even with pooled testing.^12^ While this resource intensive sampling strategy is useful for capturing the overall presence of a disease, alternative environmental sampling can serve as a warning sign of early-stage disease presence in a community prior to symptomatic patients testing positive.^13,14^

One example of passive viral sensing is testing for SARS-CoV-2 in community wastewater plants.^13,15–18^ However, wastewater surveillance methods suffer from a fixed, coarse granularity since sampling happens far downstream from the individual source. Leveraging multiple environmental sampling techniques through additional infrastructural media, such as public transit, can make viral monitoring more robust. Wastewater sampling has been explored for commercial aircraft and cruise ships,^19^ but these approaches cannot be extended to public transport vehicles without wastewater management facilities. Public transit such as buses, light rails, and trains may be valuable targets for surveillance sampling, since they are linked to the population’s geospatial mobility. The United Nations estimated *>*50% global population lives in urban centers,^20^ such as Seattle, where nearly 50% of urban commuters use public transit.^21^

Viral particles expelled from the respiratory system of an infected individual can circulate through the air into Heating, Ventilation, and Air Conditioning (HVAC) systems, and have been detected in air filters in hospitals treating infected individuals,^22–26^ suggesting a similar approach for public transit. Prior work has examined risk of transmission for passengers on buses, trains, and airplanes at local, national, and international scale; ^27–39^ however, these reports have not leveraged public transportation for community monitoring. One potential reason for this is the cost and time associated with known sampling methods with adequate Limits of Detection (LOD) to sense the low number of copies of virus expected in filters without employing active systems of collection, such as environmental swabbing or vacuum-like Personal Environmental Monitor (PEM) equipment.^40^ Rapid and inexpensive RNA extraction methods have detected 10-20 copies/reaction, which may be above the viral copies recovered from passive HVAC systems in non-concentrated settings outside of hospitals.^41^ Additionally, virus particles can remain viable for 7 days on porous surfaces, like air filters, and 3 days on non-porous surfaces, like metal hand-grips.^42,43^ Therefore, air filters may accumulate and maintain virus over a longer time than swabbed surfaces, capturing data from more individuals with a single sample, enabling pooled testing.

Here, we explore the feasibility of passive surveillance sampling in public buses by installing fabric sensors in vehicle air filtration systems. We demonstrate that sensitive methods of detection can be used to detect small copy numbers from samples collected from bus filters, using viral lysis, RNA extraction, and RNA detection via reverse transcription quantitative polymerase chain reaction (RT-qPCR) in a combination not proven in prior literature. Although prior work has demonstrated species extraction from building air filters,^26,44^ our method is a novel, passive strategy for local SARS-CoV-2 surveillance monitoring in urban transit, which shows high analytical sensitivity and specificity for low concentration environments. We evaluated this in-house method in samples collected from actively circulating buses to demonstrate the detection of SARS-CoV-2 RNA in real-world environments, and we present herein an analysis of how this method can relate to citywide cases for future disease monitoring use cases.

## Methods

### Sample Collection from Public Buses

Between August 2020 and March 2021, environmental samples were collected from 15 actively deployed buses in the Seattle King County Metro fleet (Figure 1A). Bus selection was narrowed down to the main bus depot that serviced the Downtown Seattle area, which has the highest ridership. Individual buses were selected to be sampled via a convenience sampling approach based on which buses could be made available at the depot on a regular basis between 7:00-9:00 AM for sample retrieval.

**Figure 1:**
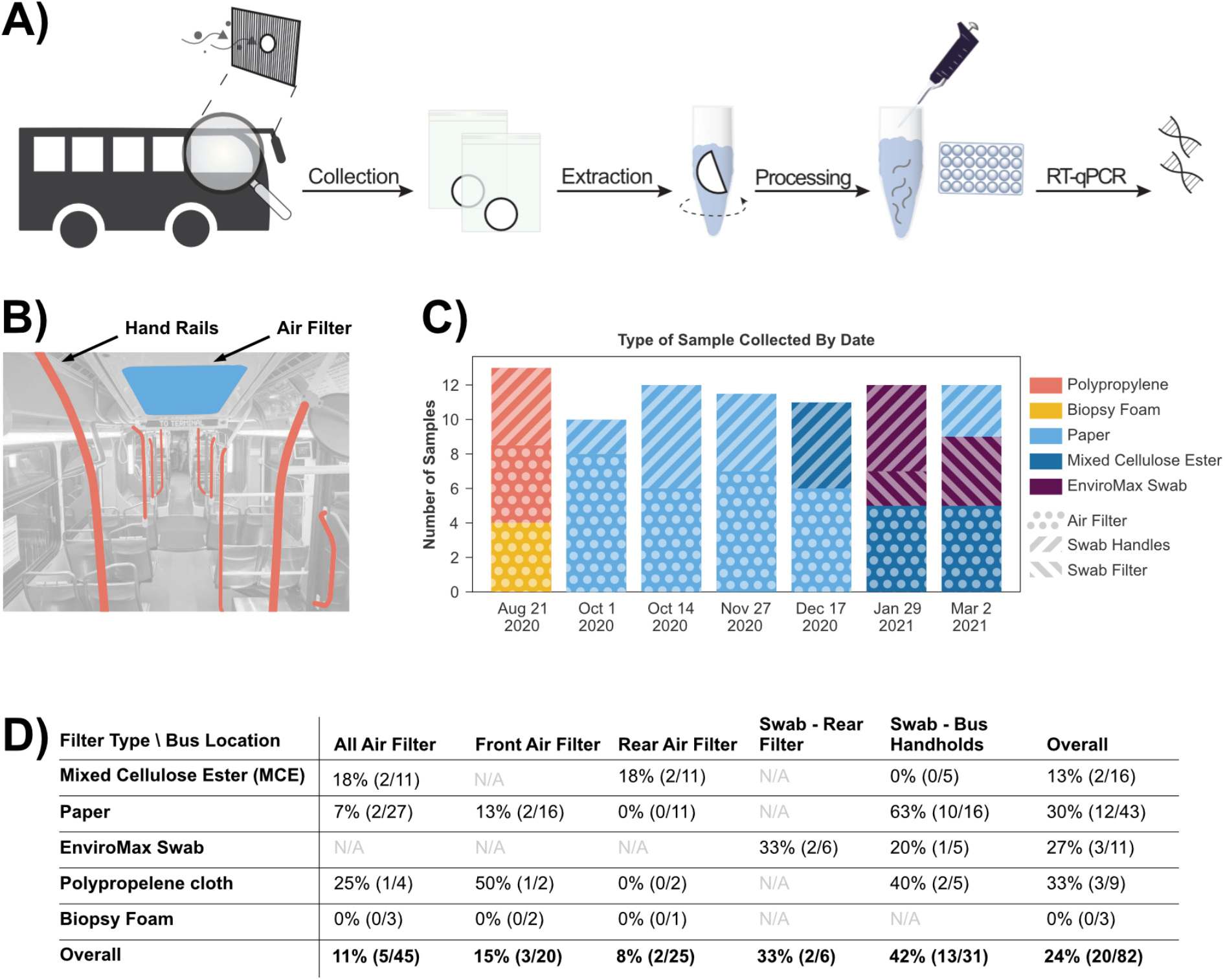
Detection of the samples collected from the metro bus using our in-house extraction protocol. (**A**) Workflow for passive sensing SARS-CoV-2 RNA including sample collection, sample transfer from papers or swabs, RNA extraction, and RT-qPCR for detection. (**B**) Sampling occurred via two methods in different areas of the bus. We collected supplementary pre-filters after more than 7 days of being installed inside the HVAC systems of actively-used metro buses (blue). We also swabbed commonly-touched surfaces on the bus (red). (**C**) Sample types and collection methods used during the course of the study. (**D**) Positivity rate breakdown by collection material and location. Sampling from both air filters as well as surfaces returned traces of SARS-CoV-2. Swabs from bus handholds made up the majority of SARS-CoV-2 detections with 42% positivity rate (13/31), while materials placed in air filters had the lowest positivity rate at 11% (5/45).

Air filters and environmental swabs were used to capture samples on buses. For air filtration testing, four different materials were tested as supplementary air filters: foam biopsy pads (22-038-221, Fisher Scientific), PolyPro fabric (25PPMB, CanvasETC), mixed cellulose ester filters (A020A025A, Thomas Scientific), and paper filters (Figure 1). Supplementary 5 cm^2^ filters were placed in front of the existing air filter in the bus HVAC system in both the back and rear of the bus. On filter collection day, four different environmental swab materials were also tested: PolyPro fabric, microporous paper separators, mixed cellulose ester filters, and EnviroMax Swabs. Swabbing was performed by running one swab across common hand-hold areas (Figure 1B in red). When EnviroMax swabs were available, a second swab of the front face of the main bus filter (Figure 1B in blue) was performed.

All bagged samples were placed in a plastic secondary container, which was wiped with bleach-based disinfectant, and transported to an approved lab facility. All procedures involving the untreated filter material were performed in a BSL2-certified Class II A2 biosafety cabinet. All types of filters that were used are shown in Figure 1C. Due to safety-related lab space and chemical SOP limits for phenol-chloroform isoamyl extraction, a maximum of n=6 buses (2 replicates for each of the two methods - filter and swab) could be tested in a single experiment.

### Detection of SARS-CoV-2 RNA in Filters

Sample extraction for testing was performed within the same day of the sample collection from metro buses. Detection of SARS-CoV-2 RNA consisted of the following steps: viral extraction and lysis, RNA isolation via phenol-chloroform isoamyl extraction, and RNA detection via RT-qPCR (Figure 1A).

### RNA Extraction and Isolation

Filters collected from buses were cut into 2-cm^2^ pieces. Two pieces, considered sample replicates, were placed into microcentrifuge tubes containing 200 µL lysis buffer (50mM EDTA pH 8.0, 250mM Tris-HCl pH 8.0, 50mM NaCl, 1% (w/v) SDS).^45^ The tubes were placed on a foam tube rack attached to a vortexer and agitated for 15 minutes, at high speed, at room temperature. After vortexing, 600 µL TRIzol was added to each tube, pipette-mixed 10 times, and then the resultant 800 µL solution was transferred into a new tube. The solutions were incubated at room temperature for 5 minutes to allow complete dissociation of viral particles into the upper media and inactivation of any potentially remaining active virus in the solution. The RNA was then isolated from protein and DNA following the standard TRIzol phase separation procedure.^46^ Precipitation of RNA was carried out by adding 1 mL 200-proof ethanol and 1 µL RNA-grade glycogen (R0551, ThermoFisher) to each tube, followed by 1-minute vortexing. Each tube was then incubated overnight at −20°C. Following overnight precipitation, the supernatant was discarded and residual ethanol was allowed to evaporate. The RNA pellet was washed following the standard TRIzol RNA wash procedure and subsequently re-suspended in 8 µL nuclease-free water.

### Detection of SARS-CoV-2 RNA Using RT-qPCR

Each 8 µL TRIzol isolation product was assayed with TaqPath 1-step RT-qPCR (A15299, ThermoFisher Scientific) in 20 µL reactions. We used probes from the CDC SARS-CoV-2 qPCR probe assay targeting two regions in the N gene, designated N1 and N2 (10006713, Integrated DNA Technologies), one for each sample replicate. To avoid cross-contamination, the reactions were loaded into non-adjacent wells in a 96-well plate on ice at a separate bench from where the RNA isolation step was performed. Wells also were covered with Parafilm between loading samples. RT-qPCR was carried out on a Quantstudio 3 (ThermoFisher) using the CDC-recommended protocol.^47^

### Positivity Determination

Positive results were determined by amplification before a specified PCR cycle threshold (CT). Raw RT-qPCR amplification data were loaded via a custom Python script (Supplementary Information) that determined CT based on a common threshold across all samples (50000 RFU) which delineated between amplified samples and non-amplified samples in control experiments. For environmental samples, a positive result for a bus was declared if one replicate from one of the sample methods from that bus had a CT *<*40.

## Results

### Results on Environmental Samples from Buses

Out of 82 samples (164 total with replicates) tested, 24% (20/82, 95% CI: 16-35) of samples tested positive for SARS-CoV-2 RNA (Figure 1D). Samples collected by the swabbing method showed the highest positivity rate at 42% (13/31, 95% CI: 25-61) while filters had the lowest positivity rate at 11% (5/45, 95% CI: 4-24). 1 replicate amplified in all positive bus samples (Table S2). This indicates that the viral particles may not be distributed evenly across the sampling material or that the sample methods may be sensing viral presence from different signal sources (i.e. riders who breathe may not touch the railing). Most positive samples had SARS-CoV-2 RNA near or below the LOD of the RT-qPCR assay (Figure S6) and thus were confirmed correct product sizes by fragment analysis (Figure S7).

We compared our TRIzol-based RNA extraction method with a more commonly used column-based RNA extraction from Qiagen. After dividing five filters in half and processing in parallel, we found a bus positivity rate of 60% (3/5) and 80% (4/5) (Table S3) in TRIzol-based and column-based methods, respectively. Interestingly, our TRIzol-based method did not yield positive results from any samples collected by EnviroMax swabs, which were positive with the column-based method. We observed black particle residues in samples using EnviroMax swabs (Figure S8), which were filtered out by the column-based extraction. These residues ended up in the RT-qPCR reactions when EnviroMax samples were extracted by the TRIzol-based method, which may have inhibited the RT-qPCR reaction. On the other hand, the column-based method displayed 0% positivity rate on all air filter samples, while the Trizol-based method detected 40% (2/5) positivity on the same air filters. We hypothesize that the debris broken from filters could interfere with the binding of SARS-CoV-2 RNA to the silica membrane in the columns or the SARS-CoV-2 RNA might remain trapped in the columns. These filters are made from mixed cellulose ester which have different surface properties and porous structures from those of polyurethane foam structures (EnviroMax swabs) which reportedly had a high release efficiency even with minimal agitation.^48^

All air filters were installed on buses for more than 7 days, and thus can represent pooled samples of all riders for the prior 7 days (Figure 2). One exception is that, for one sampling date on October 14, 2021, filters were installed and collected in one day. In one-day testing, 0 filters returned positive, indicating that one day may not be enough filter exposure time to build up a detectable viral load. However, a relatively high rate (60%) of swab samples returned positive, which may be attributable to lack of surface decontamination mid-day. Metro cleans buses nightly, and the morning sampling period for all 2-week samples occurred the morning following the decontamination, in between which no riders would have ridden the bus.

**Figure 2:**
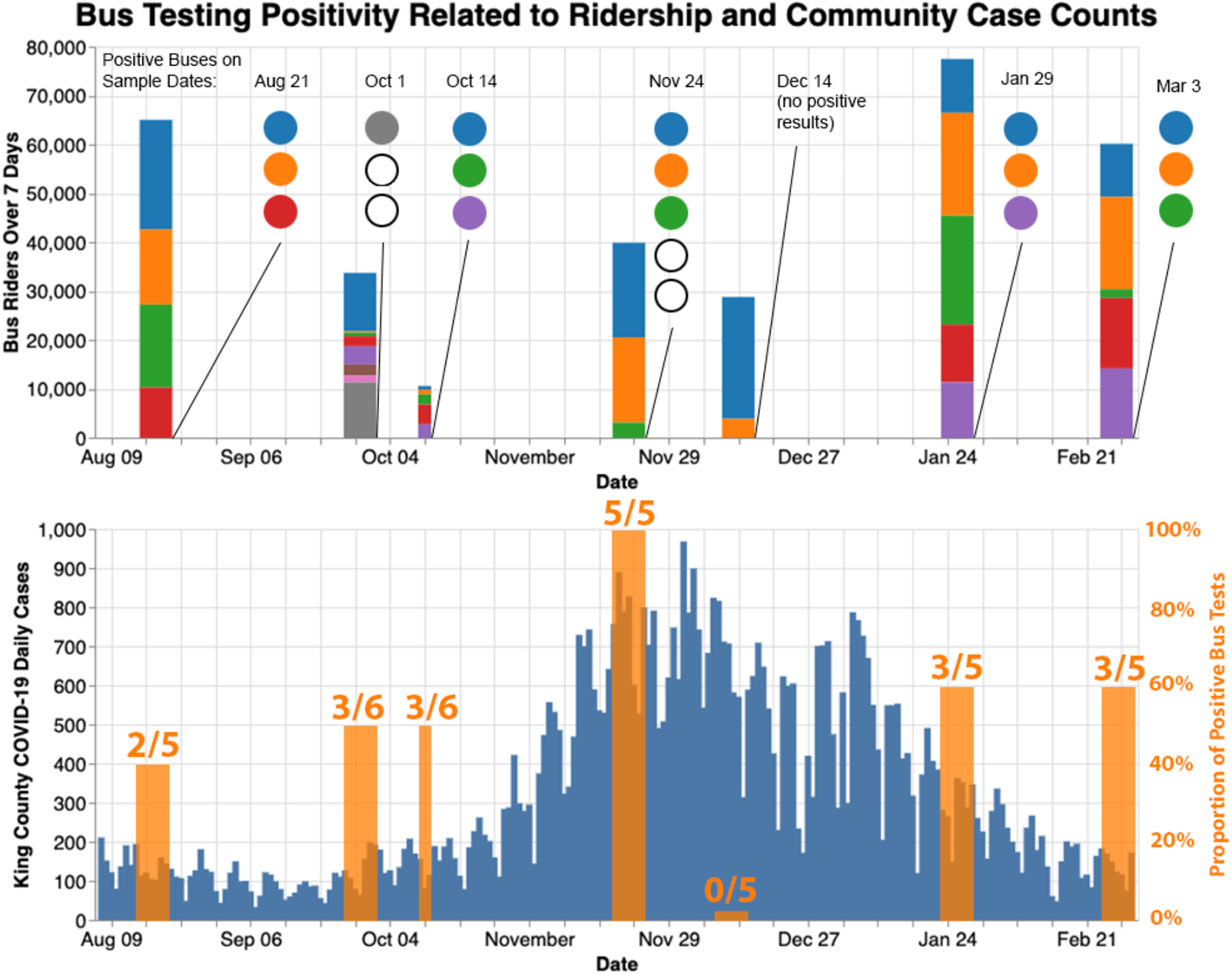
**Top** chart shows total riders per bus per 7-day period of filter installation before sampling. Each color denotes a unique bus that week. The color of associated circles denotes a positive result from that bus. An empty circle denotes a positive sample for a bus with 0 riders during that week. **Bottom** chart shows new cases of SARS-CoV-2 in King County (blue) superimposed with the proportion of buses sampled that week returning positive results (orange).

Figure 2 shows the bus testing results juxtaposed with the SARS-CoV-2 case counts and bus ridership counts for the 7 sampling period. A higher proportion of buses sampled return positive results when SARS-CoV-2 cases in King County were high. December 14th showed no positive results, although cases were high. This is likely due to the fact that only 2/5 buses sampled had any ridership during that week. October 1st and November 24th also returned positive results for buses which did not leave the station (zero riders), denoted by the empty circles in Figure 2. This may have been the result of continued viral RNA presence after more than 7 days or infected maintenance workers who entered a bus during the sample period. Metro’s ridership data was compared to bus positivity results to determine whether ridership was correlated with the positivity rate for buses, and a small Pearson’s Correlation of 0.255 was observed. This correlation may have been reduced due to infection control measures taken by King County Metro, such as required rider masking and nightly surface disinfection (Virex). Additionally, the population of metro riders from which the testing was sampled is not fully representative of the overall population from which individual testing was performed.

## Discussion

Here we show that passive infrastructure-mediated sensing of viral presence may be feasible. By leveraging a passive viral sensing method such as ours in parallel with other environmental and individual testing methods, epidemiologists could inexpensively monitor a community to identify locales of transmission and estimate case numbers within local regions. Our method may be more valuable when cases within the general community are low, leveraging the fabric sensors to passively pool respiratory droplet samples from riders temporally over the filter installation period and spatially over bus routes.

This study is limited by its sample size (n=39 total buses). Sample filters were placed and recovered manually by the research team and metro collaborators, which could be scaled by larger research teams (Table S4). False negatives may have been caused by the limited coverage of the sample media over the air vent and air currents diverting around the sample filter media. In addition, mask mandates were in effect for riders during the sample period, likely reducing the number of viral particles from infected riders landing on the filters. Considering these effects, the small viral loads (Figure S6) of some samples are unsurprising, but may fall below the typical LOD for many commercial PCR kits, including our chosen Taqpath 1-step kit.^47^ Thus, false negatives may occur due to loss in the extraction and PCR steps. We also note that our method does not necessarily identify the risk to bus riders, but rather the presence of inactive SARS-CoV-2 RNA. Viral viability tests and more frequent sampling are needed to understand the risk to riders.

Future research into scalable, sensitive viral detection for environmental samples would enhance this approach. Studies evaluating filter placement and size, as well as control experiments in simulated environments, could further validate the sensitivity of the method. City-wide deployments enabled by scalable detection methods, such as rapid diagnostic lateral flow detection for on-site detection enabled by miniaturized PCR devices^41^ or sequencers,^49^ could gather more data, enabling network analysis techniques to study probability of SARS-CoV-2 transmission on a neighborhood level. This method could be adapted and deployed to provide an early signal of community outbreak of SARS-CoV-2, or other viruses transmitted by respiratory droplets, when case counts are low in the population. This could provide a relatively inexpensive early warning system and ongoing monitoring insight into the local routes of viral transmission for current and future respiratory pathogens.

## Data Availability

Data is shared in Supplemental Information and will be provided via Github shortly.

## Acknowledgement

We thank Jon Custer and other members at King County Metro for collaboration with sample collection and sharing ridership information. We thank Karin Strauss, Mike Reddy, and other collaborators from Microsoft Research for financially supporting the research and providing excellent feedback in periodic discussions. We want to thank Dr. Georg Seelig and the Synthetic Biology Lab for allowing us to use their lab facilities. We also want to thank Dr. Scott Meschke for feedback on the manuscript. This work was supported by the University of Washington Population Health Department for the Economic Recovery grant and the Microsoft Studies in Pandemic Preparedness and AI for Health programs.

## Conflict of Interest Statement

The authors declare no competing financial interest.

## Supporting Information Available

### List of Supplementary Contents

1. Figure S1-5 Amplification curves of RT-qPCR from extraction controls, RNA standards, and bus samples.
2. Figure S6 Threshold cycles (CT) of RT-qPCR from extraction controls, RNA standards, and bus samples.
3. Table S1 PCR CTs.
4. Table S2 Bus CTs.
5. Figure S9 Fragment analysis of positive products using polyacrylamide gel electropheresis
6. Table S3 Comparison to column-based RNA detection
7. Figure S11 Extracted RNA solution from swab samples after TriZOL and Qiagen extraction
8. Table S4 Cost analysis of procedure
9. Open source code availability

## Notes

### Competing Interest Statement

The authors have declared no competing interest.

### Author Declarations

No IRB approval, as this was an environmental study involving no study participants.

## References

(1) Centers for Disease Control and Prevention, CDC Covid Data Tracker. 2020; https://covid.cdc.gov/covid-data-tracker/.

(2) World Health Organization and others, Coronavirus disease (COVID-2019) situation reports. 2020; https://www.who.int/emergencies/diseases/novel-coronavirus-2019/situation-reports/.

(3) Buonanno, G.; Morawska, L.; Stabile, L. Quantitative Assessment of the Risk of Air-borne Transmission of SARS-CoV-2 Infection: Prospective and Retrospective Applications. medRxiv 2020, 2020.06.01.20118984.

(4) Zhang, R.; Li, Y.; Zhang, A. L.; Wang, Y.; Molina, M. J. Identifying Airborne Transmission as the Dominant Route for the Spread of COVID-19. Proceedings of the National Academy of Sciences of the United States of America 2020, 117, 14857–14863.

(5) Wiersinga, W. J.; Rhodes, A.; Cheng, A. C.; Peacock, S. J.; Prescott, H. C. Pathophysiology, Transmission, Diagnosis, and Treatment of Coronavirus Disease 2019 (COVID-19): A Review. JAMA 2020,

(6) Bai, Y.; Yao, L.; Wei, T.; Tian, F.; Jin, D.-Y.; Chen, L.; Wang, M. Presumed Asymptomatic Carrier Transmission of COVID-19. JAMA 2020, 323, 1406–1407.

(7) Rothe, C. et al. Transmission of 2019-nCoV Infection from an Asymptomatic Contact in Germany. The New England Journal of Medicine 2020, 382, 970–971.

(8) Yu, P.; Zhu, J.; Zhang, Z.; Han, Y. A Familial Cluster of Infection Associated With the 2019 Novel Coronavirus Indicating Possible Person-to-Person Transmission During the Incubation Period. The Journal of Infectious Diseases 2020, 221, 1757–1761.

(9) Hu, Z.; Song, C.; Xu, C.; Jin, G.; Chen, Y.; Xu, X.; Ma, H.; Chen, W.; Lin, Y.; Zheng, Y.; Wang, J.; Hu, Z.; Yi, Y.; Shen, H. Clinical Characteristics of 24 Asymptomatic Infections with COVID-19 Screened among Close Contacts in Nanjing, China. Science China. Life Sciences 2020, 63, 706–711.

(10) Yu, X.; Yang, R. COVID-19 Transmission through Asymptomatic Carriers Is a Challenge to Containment. Influenza and Other Respiratory Viruses 2020, 14, 474–475.

(11) Huff, H. V.; Singh, A. Asymptomatic Transmission during the COVID-19 Pandemic and Implications for Public Health Strategies. Clinical Infectious Diseases: An Official Publication of the Infectious Diseases Society of America 2020,

(12) Augenblick, N.; Kolstad, J. T.; Obermeyer, Z.; Wang, A. Group testing in a pandemic: The role of frequent testing, correlated risk, and machine learning; 2020.

(13) Daughton, C. G. Wastewater Surveillance for Population-Wide Covid-19: The Present and Future. Science of The Total Environment 2020, 736, 139631.

(14) World Health Organization, Status of Environmental Surveillance for SARS-CoV-2 Virus. 2020.

(15) Wurtzer, S.; Marechal, V.; Mouchel, J.-M.; Maday, Y.; Teyssou, R.; Richard, E.; Almayrac, J. L.; Moulin, L. Evaluation of Lockdown Impact on SARS-CoV-2 Dynamics through Viral Genome Quantification in Paris Wastewaters. medRxiv 2020, 2020.04.12.20062679.

(16) Wu, F. et al. SARS-CoV-2 Titers in Wastewater Are Higher than Expected from Clinically Confirmed Cases. medRxiv 2020, 2020.04.05.20051540.

(17) Peccia, J.; Zulli, A.; Brackney, D. E.; Grubaugh, N. D.; Kaplan, E. H.; Casanovas-Massana, A.; Ko, A. I.; Malik, A. A.; Wang, D.; Wang, M.; Weinberger, D. M.; Omer, S. B. SARS-CoV-2 RNA Concentrations in Primary Municipal Sewage Sludge as a Leading Indicator of COVID-19 Outbreak Dynamics. medRxiv 2020, 2020.05.19.20105999.

(18) Medema, G.; Heijnen, L.; Elsinga, G.; Italiaander, R.; Brouwer, A. Presence of SARS-Coronavirus-2 RNA in sewage and correlation with reported COVID-19 prevalence in the early stage of the epidemic in the Netherlands. Environmental Science & Technology Letters 2020, 7, 511–516.

(19) Ahmed, W. et al. Detection of SARS-CoV-2 RNA in Commercial Passenger Aircraft and Cruise Ship Wastewater: A Surveillance Tool for Assessing the Presence of COVID-19 Infected Travellers. 27.

(20) Neiderud, C.-J. How Urbanization Affects the Epidemiology of Emerging Infectious Diseases. Infection Ecology & Epidemiology 2015, 5.

(21) of Transportation, S. D. 2019 Seattle Center City Commute Mode Split Survey. https://www.commuteseattle.com/wp-content/uploads/2020/09/2019-Mode-Split-Final-Report.pdf, 2019; Accessed: 2021-05-05.

(22) Kim, S.-H.; Chang, S. Y.; Sung, M.; Park, J. H.; Bin Kim, H.; Lee, H.; Choi, J.-P.; Choi, W. S.; Min, J.-Y. Extensive Viable Middle East Respiratory Syndrome (MERS) Coronavirus Contamination in Air and Surrounding Environment in MERS Isolation Wards. Clinical Infectious Diseases: An Official Publication of the Infectious Diseases Society of America 2016, 63, 363–369.

(23) Ong, S. W. X.; Tan, Y. K.; Chia, P. Y.; Lee, T. H.; Ng, O. T.; Wong, M. S. Y.; Marimuthu, K. Air, Surface Environmental, and Personal Protective Equipment Contamination by Severe Acute Respiratory Syndrome Coronavirus 2 (SARS-CoV-2) From a Symptomatic Patient. JAMA 2020,

(24) Guo, Z.-D. et al. Aerosol and Surface Distribution of Severe Acute Respiratory Syndrome Coronavirus 2 in Hospital Wards, Wuhan, China, 2020. Emerging Infectious Diseases 2020, 26.

(25) Chirico, F.; Sacco, A.; Bragazzi, N. L.; Magnavita, N. Can Air-Conditioning Systems Contribute to the Spread of SARS/MERS/COVID-19 Infection? Insights from a Rapid Review of the Literature. International Journal of Environmental Research and Public Health 2020, 17.

(26) Horve, P. F.; Dietz, L.; Fretz, M.; Constant, D. A.; Wilkes, A.; Townes, J. M.; Martindale, R. G.; Messer, W. B.; Wymelenberg, K. V. D. Identification of SARS-CoV-2 RNA in Healthcare Heating, Ventilation, and Air Conditioning Units. medRxiv 2020, 2020.06.26.20141085.

(27) Hu, M.; Lin, H.; Wang, J.; Xu, C.; Tatem, A. J.; Meng, B.; Zhang, X.; Liu, Y.; Wang, P.; Wu, G.; Xie, H.; Lai, S. The Risk of COVID-19 Transmission in Train Passengers: An Epidemiological and Modelling Study. Clinical Infectious Diseases: An Official Publication of the Infectious Diseases Society of America 2020,

(28) Zheng, R.; Xu, Y.; Wang, W.; Ning, G.; Bi, Y. Spatial Transmission of COVID-19 via Public and Private Transportation in China. Travel Medicine and Infectious Disease 2020, 34, 101626.

(29) Zhao, S.; Zhuang, Z.; Ran, J.; Lin, J.; Yang, G.; Yang, L.; He, D. The Association between Domestic Train Transportation and Novel Coronavirus (2019-nCoV) Outbreak in China from 2019 to 2020: A Data-Driven Correlational Report. Travel Medicine and Infectious Disease 2020, 33, 101568.

(30) Browne, A.; Ahmad, S. S.-O.; Beck, C. R.; Nguyen-Van-Tam, J. S. The Roles of Transportation and Transportation Hubs in the Propagation of Influenza and Coronaviruses: A Systematic Review. Journal of Travel Medicine 2016, 23.

(31) Hoehl, S.; Karaca, O.; Kohmer, N.; Westhaus, S.; Graf, J.; Goetsch, U.; Ciesek, S. Assessment of SARS-CoV-2 Transmission on an International Flight and Among a Tourist Group. JAMA Network Open 2020, 3, e2018044–e2018044.

(32) Shen, Y. et al. Community Outbreak Investigation of SARS-CoV-2 Transmission Among Bus Riders in Eastern China. JAMA Internal Medicine 2020,

(33) Luo, L. et al. Contact Settings and Risk for Transmission in 3410 Close Contacts of Patients With COVID-19 in Guangzhou, China. Annals of Internal Medicine 2020,

(34) Bae, S. H.; Shin, H.; Koo, H.-Y.; Lee, S. W.; Yang, J. M.; Yon, D. K. Early Release - Asymptomatic Transmission of SARS-CoV-2 on Evacuation Flight - Volume 26, Number 11—November 2020 - Emerging Infectious Diseases Journal - CDC. 2020,

(35) Yang, N.; Shen, Y.; Shi, C.; Ma, A. H. Y.; Zhang, X.; Jian, X.; Wang, L.; Shi, J.; Wu, C.; Li, G.; Fu, Y.; Wang, K.; Lu, M.; Qian, G. In-Flight Transmission Cluster of COVID-19: A Retrospective Case Series. Infectious Diseases 2020, 0, 1–11.

(36) Kong, D. et al. Clusters of 2019 Coronavirus Disease (COVID-19) Cases in Chinese Tour Groups. Transboundary and Emerging Diseases 2020, n/a.

(37) Kasper, M. R.; Geibe, J. R.; Sears, C. L.; Riegodedios, A. J.; Luse, T.; Von Thun, A. M.; McGinnis, M. B.; Olson, N.; Houskamp, D.; Fenequito, R.; Burgess, T. H.; Armstrong, A. W.; DeLong, G.; Hawkins, R. J.; Gillingham, B. L. An Outbreak of Covid-19 on an Aircraft Carrier. EJMoa2019375.

(38) Harvey, A. P.; Fuhrmeister, E. R.; Cantrell, M. E.; Pitol, A. K.; Swarthout, J. M.; Powers, J. E.; Nadimpalli, M. L.; Julian, T. R.; Pickering, A. J. Longitudinal Monitoring of SARS-CoV-2 RNA on High-Touch Surfaces in a Community Setting. 8, 168–175.

(39) Silcott, D.; Kinahan, S.; Santarpia, J.; Silcott, B.; Silcott, R.; Silcott, P.; Silcott, B.; Distelhorst, S.; Herrera, V.; Rivera, D., et al. TRANSCOM/AMC commercial aircraft cabin aerosol dispersion tests; 2020.

(40) Moreno, T.; Pint’ so, R. M.; Bosch, A.; Moreno, N.; Alastuey, A.; Minguill’on, M. C.; Anfruns-Estrada, E.; Guix, S.; Fuentes, C.; Buonanno, G.; Stabile, L.; Morawska, L.; Querol, X. Tracing Surface and Airborne SARS-CoV-2 RNA inside Public Buses and Subway Trains. 147, 106326.

(41) Panpradist, N.; Wang, Q.; Ruth, P. S.; Kotnik, J. H.; Oreskovic, A. K.; Miller, A.; Stewart, S. W.; Vrana, J.; Han, P. D.; Beck, I. A., et al. Simpler and faster Covid-19 testing: Strategies to streamline SARS-CoV-2 molecular assays. EBioMedicine 2021, 64, 103236.

(42) Aboubakr, H. A.; Sharafeldin, T. A.; Goyal, S. M. Stability of SARS-CoV-2 and other coronaviruses in the environment and on common touch surfaces and the influence of climatic conditions: A review. Transboundary and emerging diseases 2020,

(43) Matson, M. J.; Yinda, C. K.; Seifert, S. N.; Bushmaker, T.; Fischer, R. J.; van Doremalen, N.; Lloyd-Smith, J. O.; Munster, V. J. Effect of environmental conditions on SARS-CoV-2 stability in human nasal mucus and sputum. Emerging infectious diseases 2020, 26, 2276.

(44) Rosario, K.; Fierer, N.; Miller, S.; Luongo, J.; Breitbart, M. Diversity of DNA and RNA Viruses in Indoor Air As Assessed via Metagenomic Sequencing. Environmental Science & Technology 2018, 52, 1014–1027.

(45) Miura, T.; Masago, Y.; Sano, D.; Omura, T. Development of an Effective Method for Recovery of Viral Genomic RNA from Environmental Silty Sediments for Quantitative Molecular Detection. Applied and Environmental Microbiology 2011, 77, 3975–3981.

(46) Rio, D. C.; Ares, M.; Hannon, G. J.; Nilsen, T. W. Purification of RNA using TRIzol (TRI reagent). Cold Spring Harbor Protocols 2010, 2010, pdb–prot5439.

(47) for Disease Control, C.; Prevention, CDC 2019-Novel Coronavirus (2019-nCoV) Real-Time RT-PCR Diagnostic Panel. https://www.fda.gov/media/134922/download, 2020; Accessed: 2021–04-30.

(48) Panpradist, N.; Toley, B. J.; Zhang, X.; Byrnes, S.; Buser, J. R.; Englund, J. A.; Lutz, B. R. Swab sample transfer for point-of-care diagnostics: characterization of swab types and manual agitation methods. PloS one 2014, 9, e105786.

(49) Cardozo, N.; Zhang, K.; Doroschak, K.; Nguyen, A.; Siddiqui, Z.; Strauss, K.; Ceze, L.; Nivala, J. Multiplexed direct detection of barcoded protein reporters on a nanopore array. bioRxiv 2019, 837542.

