## Supplementary Figures for "Passively Sensing SARS-CoV-2 RNA in Public Transit Buses"

### Supporting Information Available

**List of Supplementary Contents**

1. Figure S1-5 Amplification curves of RT-qPCR from extraction controls, RNA standards, and bus samples.
2. Figure S6 Threshold cycles (CT) of RT-qPCR from extraction controls, RNA standards, and bus samples.
3. Table S1 PCR CTs.
4. Table S2 Bus CTs.
5. Figure S9 Fragment analysis of positive products using polyacrylamide gel electropheresis
6. Table S3 Comparison to column-based RNA detection
7. Figure S11 Extracted RNA solution from swab samples after TriZOL and Qiagen extraction
8. Table S4 Cost analysis of procedure
9. Open source code availability

### Supplementary Information

#### Testing in-house extraction protocol on contrived samples

In each RT-qPCR assay, we included a negative control of two replicates containing nuclease-free water (AAJ71786AE, Fisher Scientific) that underwent the TRIzol isolation steps in parallel to the samples. Each assay also included three positive controls containing 10, 100, or 1000 copies of purified RNA controls including the N gene regions of SARS-CoV-2 (102024, Twist Biosciences, San Francisco, CA). To test the efficiency of TRIzol isolation of RNA from viral envelopes, the TRIzol extraction and RT-qPCR method was performed on AccuPlex enveloped RNA reference material (0505-0126, Seracare, Milford, MA), and results are displayed in Figure S1 below.

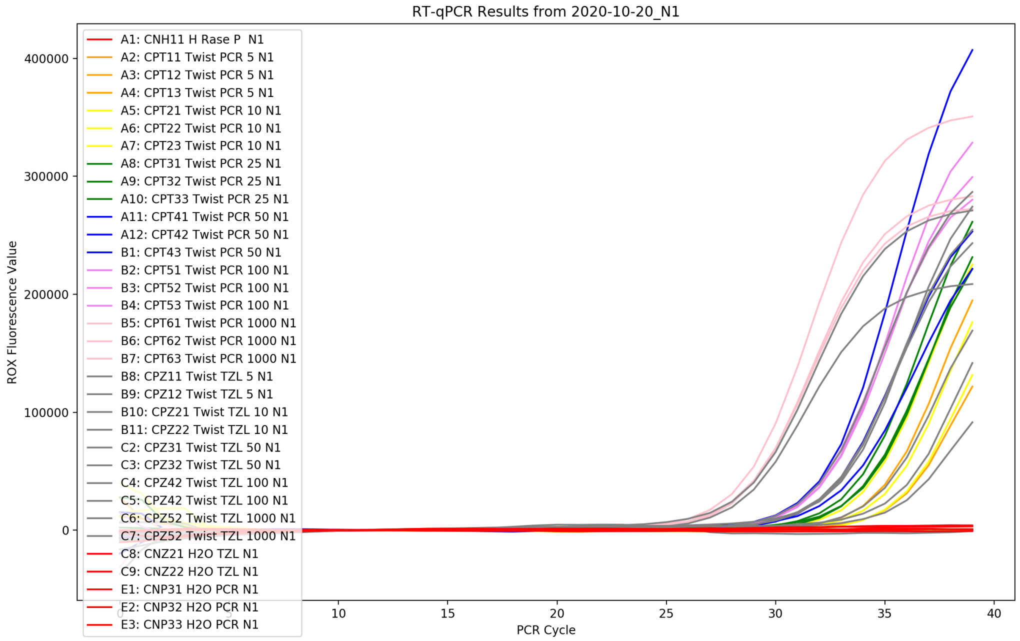


Figure S1: Controls PCR curves for N1 gene.

**Legend Key**

For Figures S1-S5:

Color, Well Number (96-well plate): Unique ID, Sample Description (including bus vehicle ID number and sample location or control type), Gene Target.

Examples:

- A4 = Column A, Row 4 of a 96-well PCR plate
- BFN12 = Bus sample, Front bus air filter, (N) Mixed Cellulose Ester filter material, 1st sample from that bus, 2nd replicate from that sample
- 4552 = Bus vehicle ID 4552
- CP12 = Control, Positive, 1st control sample, 2nd replicate of that control

Key:

- G = Polypropylene Fabric
- F = Biopsy Foam
- P = Paper
- N = Mixed Cellulose Ester
- E = EnviroMax Swab
- S = Swab Rails
- BS = Swab filter
- B, F, FS, BF = Air filter sample

#### PCR curves from bus testing

The method was also validated on environmental samples collected from buses. The procedure detected RNA from SARS-CoV-2 using both the air filter and the swab method. The results can be seen below in Figures S2-S5.

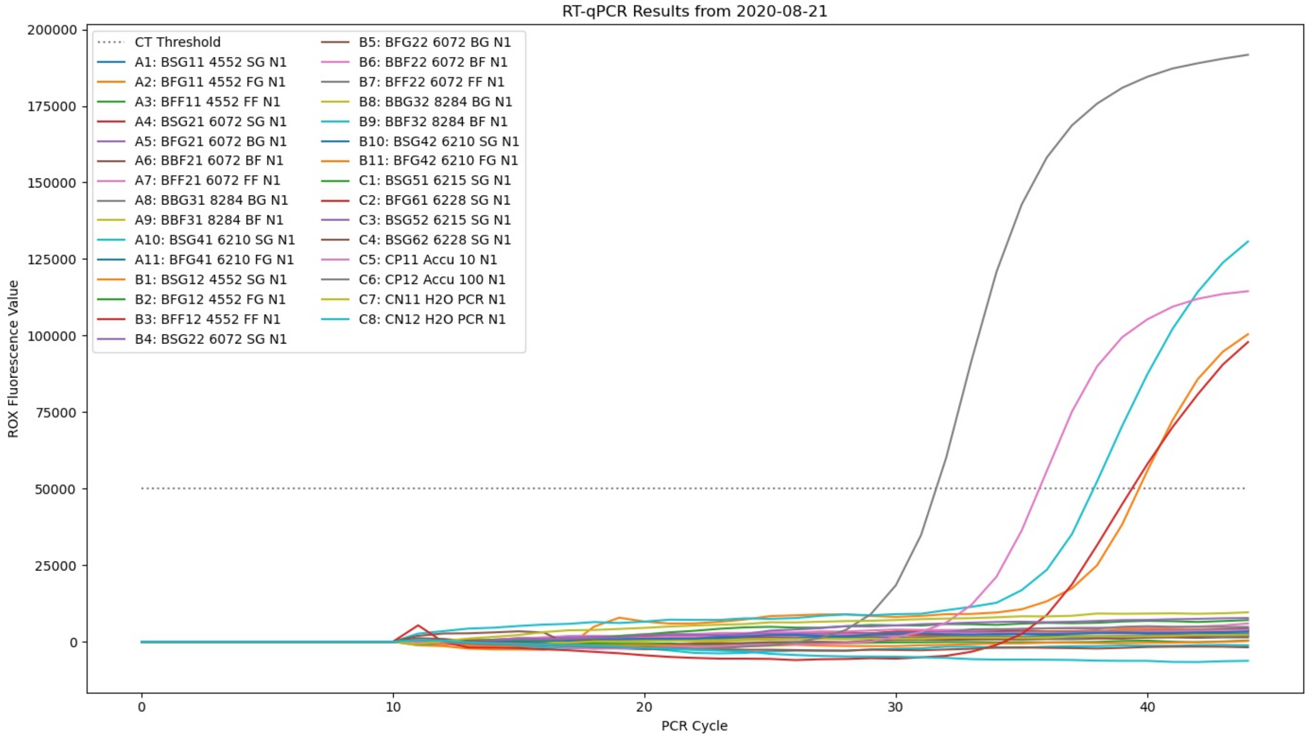


Figure S2: Bus PCRs from first positive test result in Aug 2020.

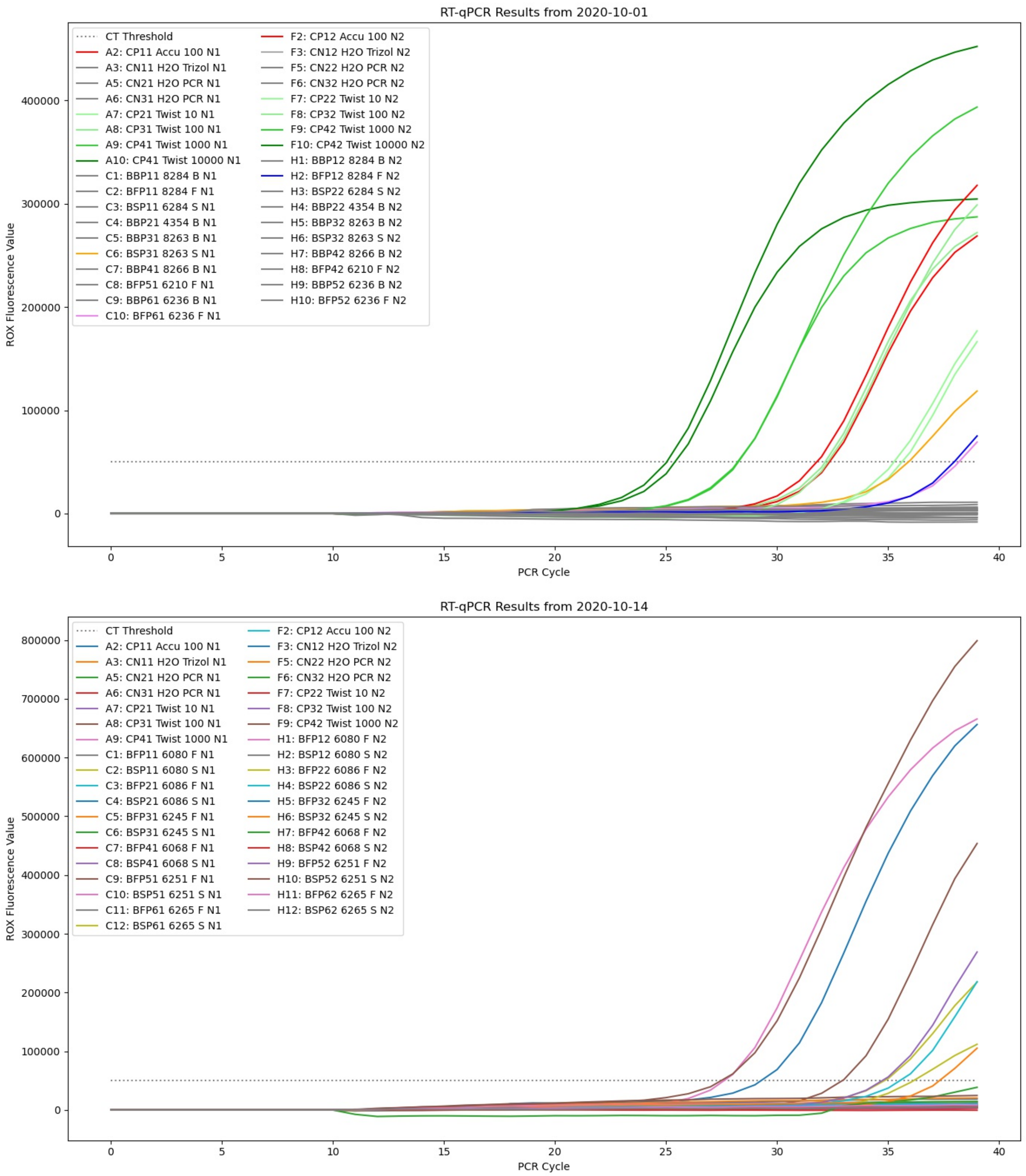


Figure S3: Bus PCRs from collection dates Oct 2020.

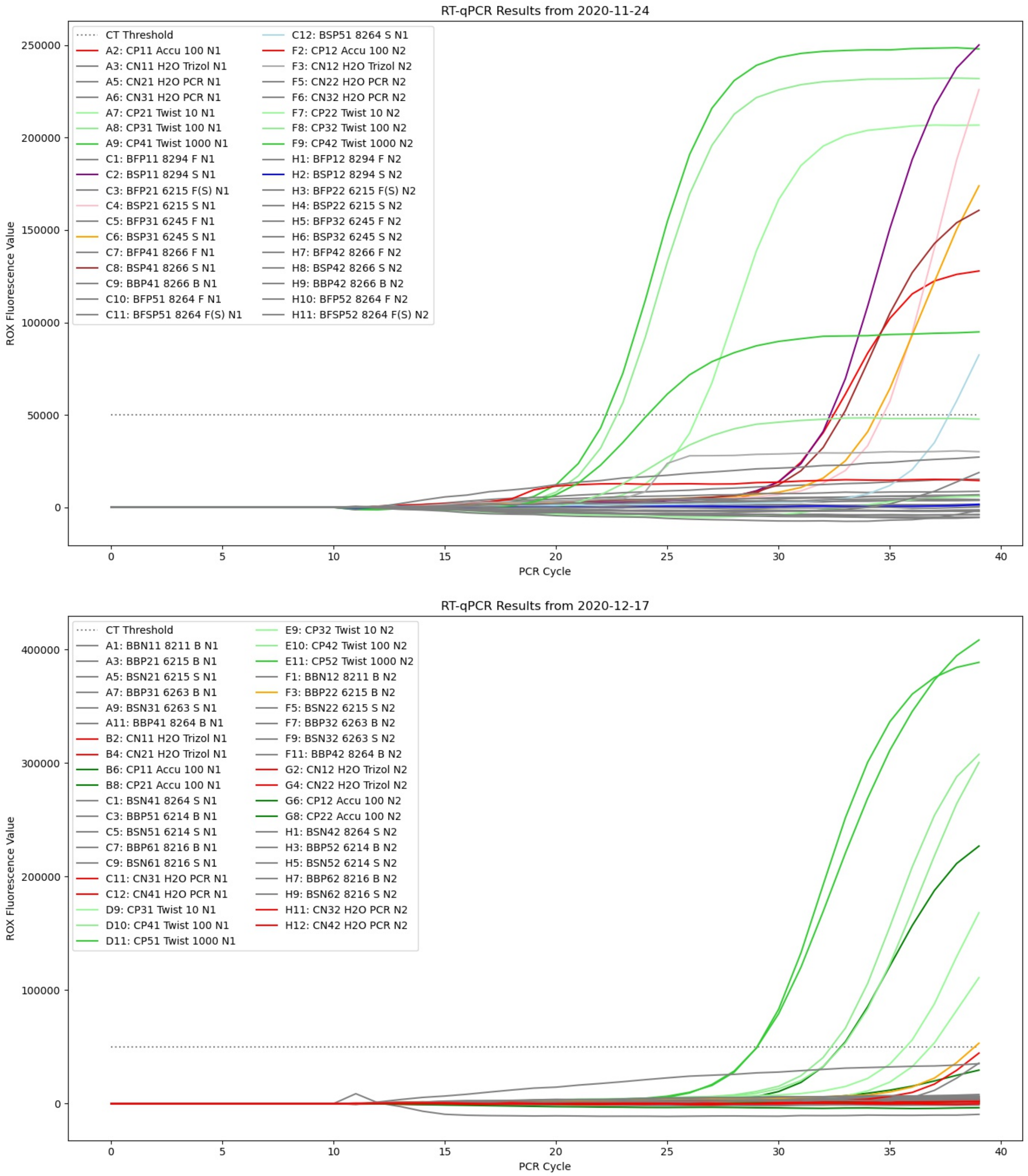


Figure S4: Bus PCRs from collection dates Nov-Dec 2020.

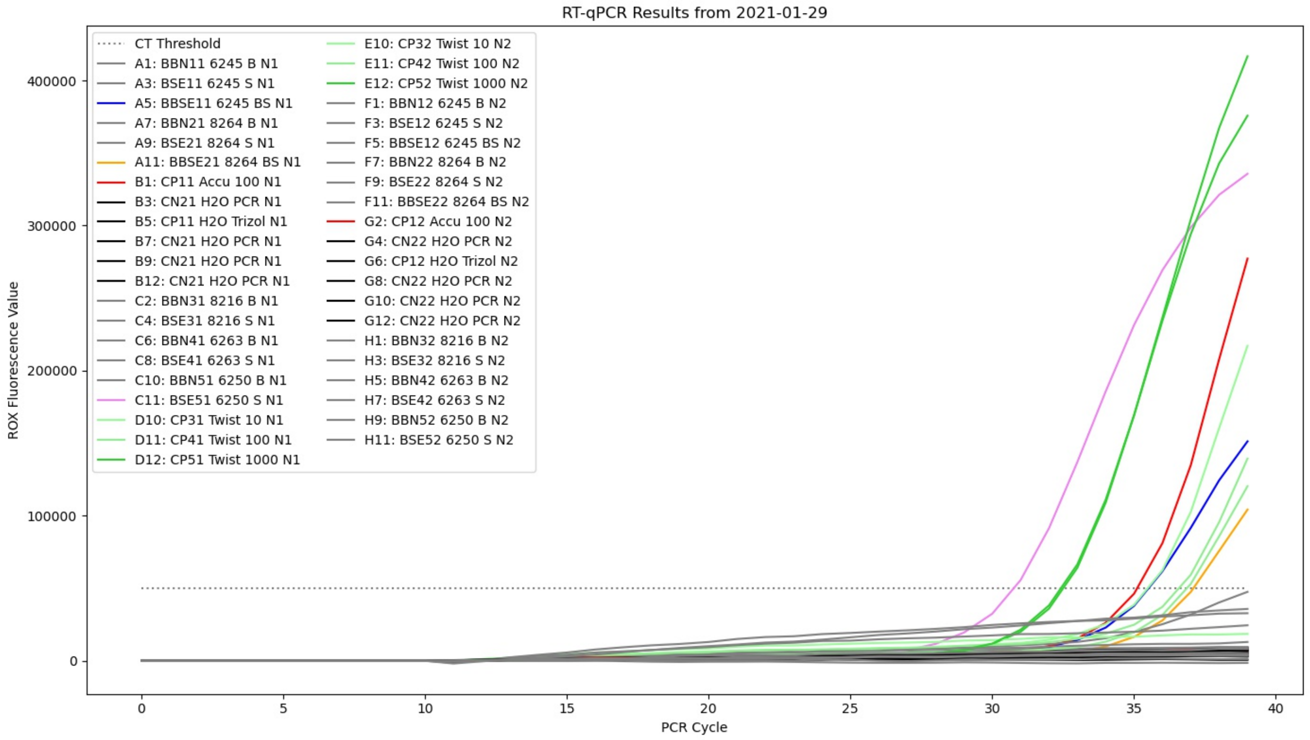


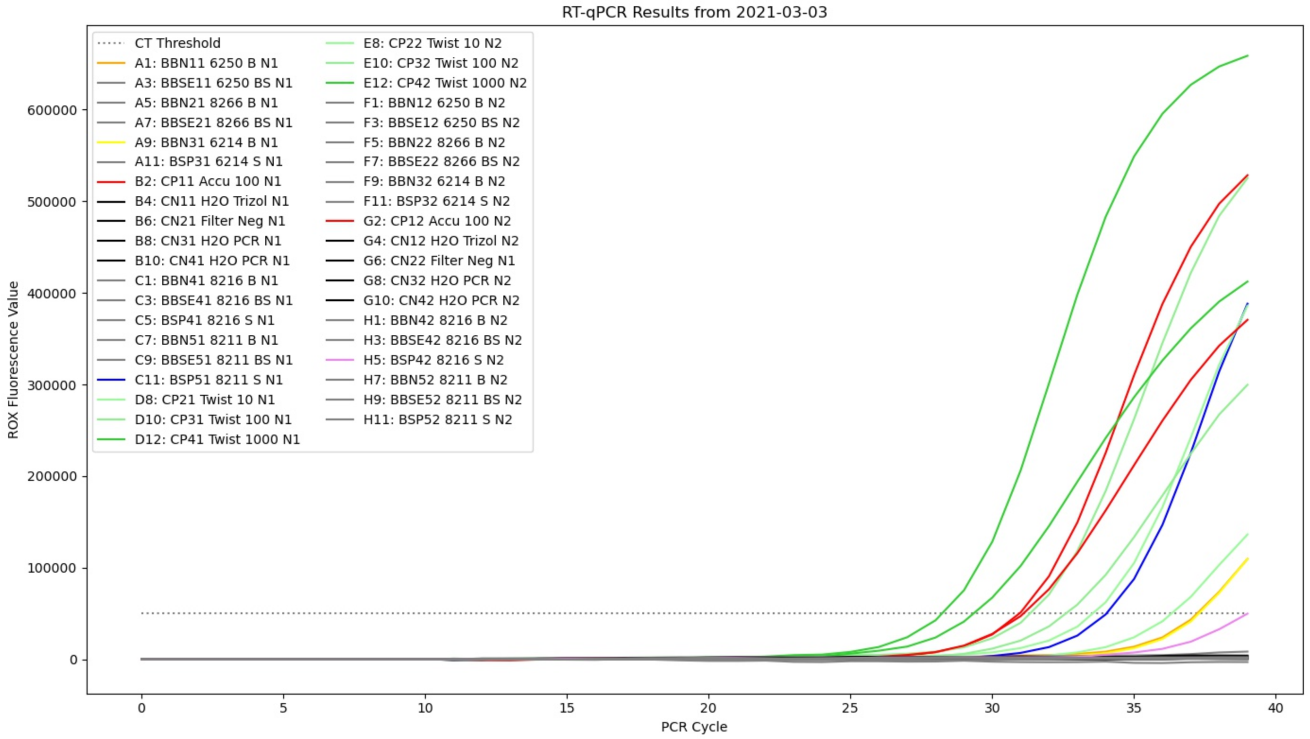


Figure S5: Bus PCRs from collection dates in Jan-March 2021.

#### qPCR Standard Curve

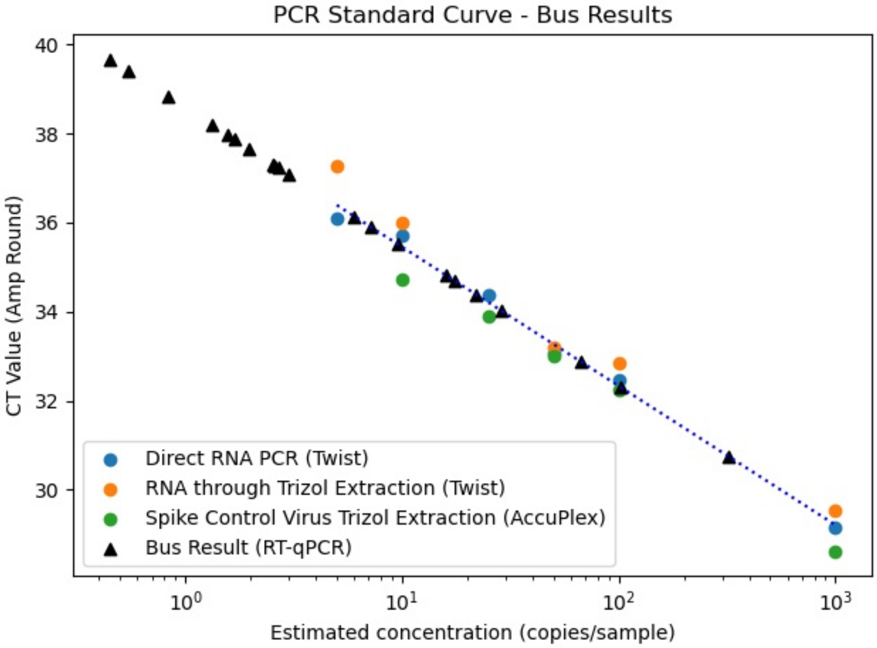


Figure S6: Creating a standard curve from the controls samples and fitting the CT results from bus runs onto that standard curve reveals most of the bus results contained a small estimated copy number of below 10 viral cells per 8 uL of extracted sample.

#### Table of results from controls testing

Table S1: CT results for controls. All controls above 10 copies/sample amplified in our testing. For copy numbers below the LOD of the PCR reagents (10 copies/sample), amplification still occurred intermittently.

| Solution | Sol Name | Copies | CT Ave | CT 1 Res | CT 2 Res | CT 3 Res |
|---|---|---|---|---|---|---|
| Direct RNA PCR (Twist) | RNA PCR | 5 | 36.10 | no amp | 35.4 | 36.79 |
| Direct RNA PCR (Twist) | RNA PCR | 10 | 35.72 | 35.8 | 34.68 | 36.68 |
| Direct RNA PCR (Twist) | RNA PCR | 25 | 34.37 | 34.47 | 34.08 | 34.55 |
| Direct RNA PCR (Twist) | RNA PCR | 50 | 33.07 | 33.17 | 33.77 | 32.27 |
| Direct RNA PCR (Twist) | RNA PCR | 100 | 32.46 | 32.36 | 32.49 | 32.52 |
| Direct RNA PCR (Twist) | RNA PCR | 1000 | 29.14 | 28.83 | 29.32 | 29.28 |
| RNA through Trizol Extraction (Twist) | RNA TZL | 5 | 37.28 | 37.28 | no amp | N/A |
| RNA through Trizol Extraction (Twist) | RNA TZL | 10 | 36.00 | 36.44 | 35.55 | N/A |
| RNA through Trizol Extraction (Twist) | RNA TZL | 50 | 33.20 | 33.23 | 33.17 | N/A |
| RNA through Trizol Extraction (Twist) | RNA TZL | 100 | 32.85 | 32.37 | 33.33 | N/A |
| RNA through Trizol Extraction (Twist) | RNA TZL | 1000 | 29.52 | 29.37 | 29.66 | N/A |
| Spike Control Virus Trizol Extraction (AccuPlex) | Accu TZL | 10 | 34.71 | 33.32 | 35.6 | 35.22 |
| Spike Control Virus Trizol Extraction (AccuPlex) | Accu TZL | 25 | 33.88 | 34.04 | 32.72 | 34.87 |
| Spike Control Virus Trizol Extraction (AccuPlex) | Accu TZL | 50 | 33.00 | 32.4 | 33.38 | 33.21 |
| Spike Control Virus Trizol Extraction (AccuPlex) | Accu TZL | 100 | 32.23 | 33.32 | 31.56 | 31.82 |
| Spike Control Virus Trizol Extraction (AccuPlex) | Accu TZL | 1000 | 28.62 | 28.14 | 29 | 28.73 |

#### Table of results from bus testing

Table S2: Full set of results from buses tested at King County Metro. All buses, replicates, and samples are recorded, alongside whether or not they amplified in PCR testing and also their CT values.

| Date Collected | Bus Number | Filter Location | Filter Material | 1 Result | 2 Result | CT @ 50000 |
|---|---|---|---|---|---|---|
| | | | | N1 | N1 | All N1 gen |
| 8/21/20 | 4552 | Swab | Gram | No | No | |
| | 4552 | Front | Gram | Yes | No | 39.67 |
| | 4552 | Front | Foam | No | No | |
| | 6072 | Swab | Gram | No | No | |
| | 6072 | Back | Gram | No | No | |
| | 6072 | Front | Foam | No | No | |
| | 8284 | Back | Gram | No | No | |
| | 8284 | Back | Foam | No | No | |
| | 6210 | Swab | Gram | Yes | No | 37.86 |
| | 6210 | Front | Gram | No | No | |
| | 6215 | Swab | Gram | No | No | |
| | 6228 | Swab | Gram | No | Yes | 39.39 |
| pos % (all bus) | 6 | 3 | 50.0% | | | |
| pos % (bus filters) | 4 | 1 | 25.0% | | | |
| | | | | N1 | N2 | N1 and N2 |
| 10/1/20 | 8284 | Back | Paper | No | No | |
| | 8284 | Front | Paper | No | Yes | 37.97 |
| | 8284 | Swab | Paper | No | No | |
| | 4354 | Back | Paper | No | No | |
| | 8263 | Back | Paper | No | No | |
| | 8263 | Swab | Paper | Yes | No | 35.91 |
| | 8266 | Back | Paper | No | No | |
| | 8266 | Front | Paper | No | No | |
| | 6210 | Front | Paper | No | No | |
| | 6236 | Back | Paper | No | No | |
| | 6236 | Front | Paper | Yes | No | 38.18 |
| pos % (all bus) | 6 | 3 | 50.0% | | | |
| pos % (bus filters) | 6 | 2 | 33.3% | | | |
| | | | | N1 | N2 | N1 and N2 |
| 10/14/20 | 6080 | Front | Paper | No | No | |
| (1 day - AM) | 6080 | Swab | Paper | Yes | No | 34.82 |
| | 6086 | Front | Paper | No | No | |
| | 6086 | Swab | Paper | No | No | |
| | 6245 | Front | Paper | No | No | |
| | 6245 | Swab | Paper | No | Yes | |
| | 6068 | Front | Paper | No | No | |
| | 6068 | Swab | Paper | No | No | |
| | 6251 | Front | Paper | No | No | |
| | 6251 | Swab | Paper | No | No | |
| | 6265 | Front | Paper | No | No | |
| | 6265 | Swab | Paper | Yes | No | 36.14 |
| pos % (all bus) | 6 | 3 | 50.0% | | | |
| pos % (bus filters) | 6 | 0 | 0.0% | | | |
| | | | | N1 | N2 | N1 and N2 |
| 11/24/20 | 8294 | Front | Paper | No | No | |
| 5 weeks | 8294 | Swab | Paper | Yes | No | 32.31 |
| | 6215 | Front (smal | Paper | No | No | |
| | 6215 | Swab | Paper | Yes | No | 34.69 |
| | 6245 | Front | Paper | No | No | |
| | 6245 | Swab | Paper | Yes | No | 34.39 |
| | 8266 | Front | Paper | No | No | |
| | 8266 | Swab | Paper | Yes | No | 32.87 |
| | 8266 | Back | Paper | No | No | |
| | 8264 | Front | Paper | No | No | |
| | 8264 | Front (smal | Paper | No | No | |
| | 8264 | Swab | Paper | Yes | N/A | 37.66 |
| pos % (all bus) | 5 | 5 | 100.0% | | | |
| pos % (bus filters) | 5 | 0 | 0.0% | | | |

| Date Collected | Bus Number | Filter Location | Filter Material | 1 Result | 2 Result | CT @ 50000 |
|---|---|---|---|---|---|---|
| | | | | N1 | N2 | |
| 12/16/20 | 8211 | Back | Nitro | No | No | |
| | 6215 | Back | Paper | No | No | |
| | 6215 | Swab | Nitro | No | No | |
| | 6263 | Back | Paper | No | No | |
| | 6263 | Swab | Nitro | No | No | |
| | 8264 | Back | Paper | No | No | |
| | 8264 | Swab | Nitro | No | No | |
| | 6214 | Back | Paper | No | No | |
| | 6214 | Swab | Nitro | No | No | |
| | 8216 | Back | Paper | No | No | |
| | 8216 | Swab | Nitro | No | No | |
| pos % (all bus) | 6 | 0 | 0.0% | | | |
| pos % (bus filters) | 6 | 0 | 0.0% | | | |
| pos % (bus- used) | 2 | 0 | 0.0% | | | |
| | | | | N1 | N2 | |
| 1/29/21 | 6245 | Back | Nitro | No | No | |
| | 6245 | Swab | EMax | No | No | |
| | 6245 | Back Swab | EMax | Yes | No | 35.51 |
| | 8264 | Back | Nitro | No | No | |
| | 8264 | Swab | EMax | No | No | |
| | 8264 | Back Swab | EMax | Yes | No | 37.09 |
| | 8216 | Back | Nitro | No | No | |
| | 8216 | Swab | EMax | No | No | |
| | 6263 | Back | Nitro | No | No | |
| | 6263 | Swab | EMax | No | No | |
| | 6250 | Back | Nitro | No | No | |
| | 6250 | Swab | EMax | Yes | No | 32.42 |
| pos % (all bus) | 5 | 3 | 60.0% | | | |
| pos % (bus filters) | 5 | 0 | 0.0% | | | |
| | | | | N1 | N2 | |
| 3/2/21 | 6250 | Back | Nitro | Yes | No | 37.22 |
| | 6250 | Back Swab | EMax | No | No | |
| | 8266 | Back | Nitro | No | No | |
| | 8266 | Back Swab | EMax | No | No | |
| | 6214 | Back | Nitro | Yes | No | 37.28 |
| | 6214 | Swab | Paper | No | No | |
| | 8216 | Back | Nitro | No | No | |
| | 8216 | Back Swab | EMax | No | No | |
| | 8216 | Swab | Paper | No | No | |
| | 8211 | Back | Nitro | No | No | |
| | 8211 | Back Swab | Emax | No | No | |
| | 8211 | Swab | Paper | Yes | No | 34.03 |
| pos % (all bus) | 5 | 3 | 60.0% | | | |
| pos % (bus filters) | 5 | 2 | 40.0% | | | |

#### Verification of Sample via native-PAGE

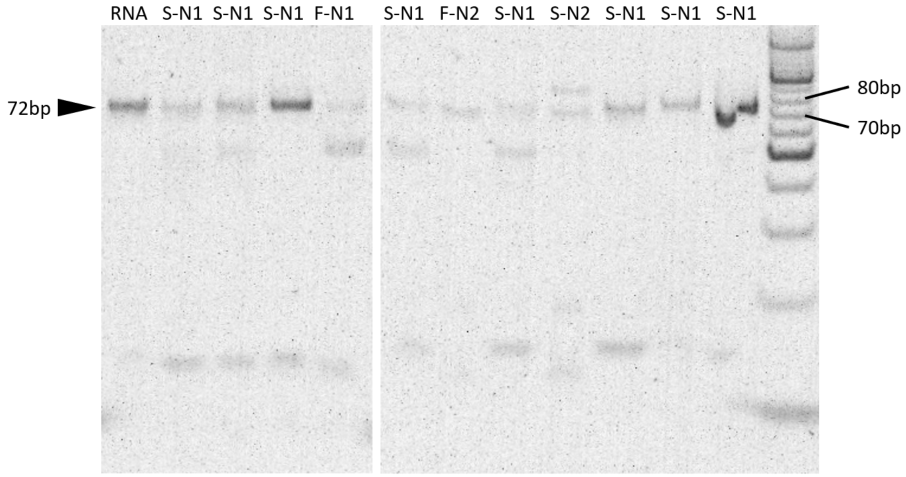


Figure S7: Randomly selected positive PCR samples were analyzed using a 12 percent polyacrylamide gel. The arrowhead indicates the expected size of the PCR product from both N1 and N2 probes. Labels differentiate between air filter and swab samples, as well as an RNA positive control. O'RangeRuler 10bp DNA Ladder (SM1313, ThermoFisher)

#### Method comparison to Qiagen column-based RNA extraction kit

Column-based extraction kits (52906, Qiagen, Hilden, Germany) were used to extract environmental samples collected on March 2nd, 2021. Each 5 µL RNA product was assayed with TaqPath 1-step RT-qPCR N1 and N2 assays and compared to its counterpart replicates processed by TRIzol extraction (Figure S3).

Table S3: A comparison was performed on 3/2/21 to understand how the results of our method compared to the silica-based extraction method. Based on these limited results, it appears that the sediments introduced by swab samples may interfere with precipitation and/or amplification of SARS-CoV-2 RNA using this method; whereas samples with very high CT (and thus low copy numbers) were not detected using Qiagen kits, indicating that this method may be more sensitive to low copy numbers when the sample is relatively clean, as is the case with passive air filter sampling.

| Date Collected | Bus ID | Filter Location | Filter Medium | Result 1 | Result 2 | CT @ 50000 ROX | Amplified? | N1-1 | N1-2 | N2-1 | N2-2 |
|---|---|---|---|---|---|---|---|---|---|---|---|
| 3/2/2021 | 6250 | Back | Nitro | Yes | No | 37.22 | No | N/A | N/A | N/A | N/A |
| | 6250 | Back Swab | EMax | No | No | | Yes | 36.7 | 36.6 | 38.27 | 35.44 |
| | 8266 | Back | Nitro | No | No | | No | N/A | N/A | N/A | N/A |
| | 8266 | Back Swab | EMax | No | No | | Yes | N/A | 37.76 | N/A | 39.42 |
| | 6214 | Back | Nitro | Yes | No | 37.28 | No | N/A | N/A | N/A | N/A |
| | 6214 | Swab | Paper | No | No | | No | N/A | N/A | N/A | N/A |
| | 8216 | Back | Nitro | No | No | | No | N/A | N/A | N/A | N/A |
| | 8216 | Back Swab | EMax | No | No | | Yes | N/A | 36.96 | N/A | N/A |
| | 8216 | Swab | Paper | No | No | | No | N/A | N/A | N/A | N/A |
| | 8211 | Back | Nitro | No | No | | No | N/A | N/A | N/A | N/A |
| | 8211 | Back Swab | Emax | No | No | | Yes | N/A | N/A | N/A | 38.47 |
| | 8211 | Swab | Paper | Yes | No | 34.03 | No | N/A | N/A | N/A | N/A |

##### Extraction Contaminants

The filter swab samples were the siltiest, returning from the bus depot with the darkest brown color. The differing ways that filter methods of TRIzol-based and column-based RNA extraction interacted withthese dirt particles likely caused difference in results for the two methods. TRIzol-based extraction resulted in positive results that column-based method missed, while the column-based method resulted in more positives in especially dirty samples.

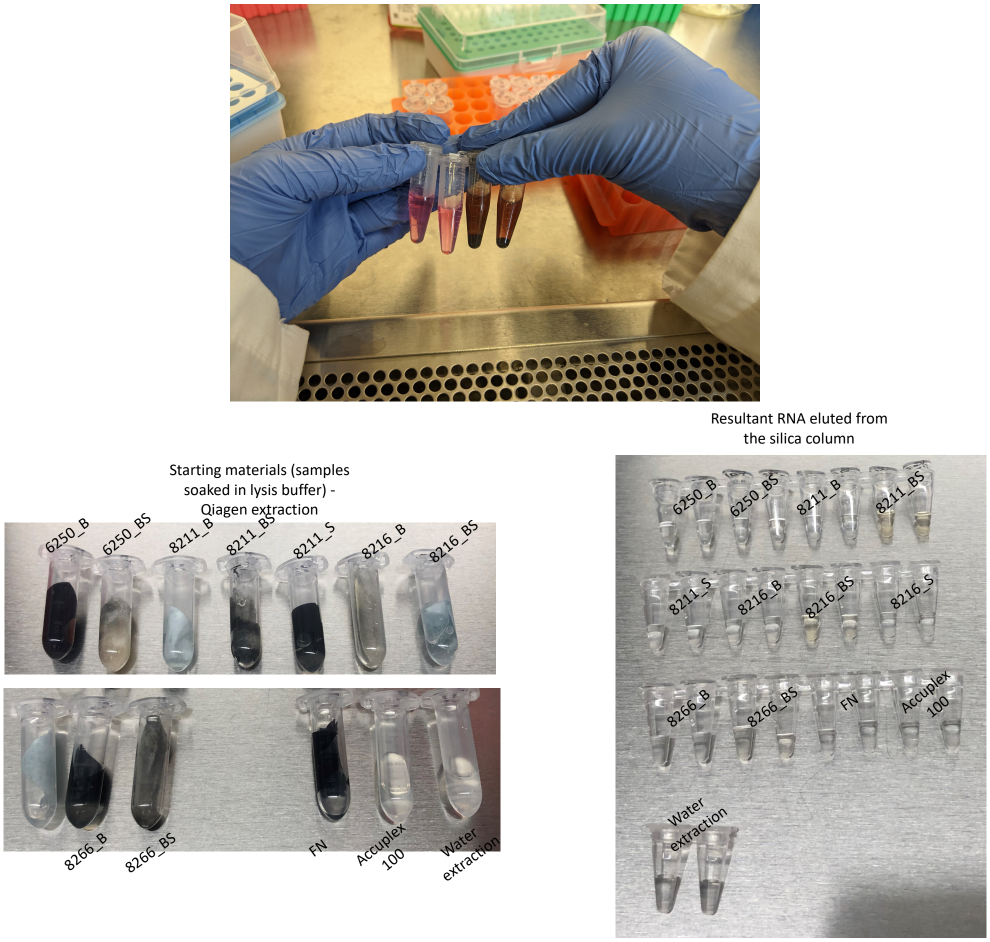


Figure S8: Swab samples' appearance after our extraction method suggests that dirt and debris could interfere with PCR results, increasing number of false negatives for especially dirty samples. **Top** Resulting tubes after extraction by TRIzol-based extraction (our method) for 2 samples from air filters (two on left) and two samples from filter swabs (two on right). The filter swab samples were especially dirty before extraction, and some of that dirt ended up in the post-extracted material. **Bottom** Resulting tubes after extraction by column-based method (Qiagen). In column-based extraction, even the dirty bus swab samples remained clear after extraction, suggesting that they filtered different content than the TRIzol-based method.

#### Cost Analysis of Method

We have shown that passive filters with minimal installation requirements are capable of capturing extremely low quantities of viral particles. However, the concentration step with TRIzol-extraction makes our procedure more time-consuming and costly per-sample than other methods, such as column-purification and amplification of nucleic acids.[47] This paper details the results of 82 individual samples separately tested across 45 buses. In theory, by combining samples from the same bus in the concentration step, or by limiting the number of locations to install filters in each bus, the effort to assay multiple buses may be reduced in. Ultimately, while our method does not include cost-prohibitive or difficult-to-obtain materials, the cost of scalability must be considered alongside the increased sensitivity when comparing to alternative detection methods (Figure S4).

Table S4: Cost breakdown of lab materials. Note that these reagent and supplies costs were based on the small scaled purchased and did not include the labor cost.

#### Total Cost

| | Total | Sampling Materials | Lab Materials |
|---|---|---|---|
| Total cost (for one test run): | **$341.66** | $58.21 | $283.45 |
| Cost / bus: | **$56.94** | $9.70 | $47.24 |

#### Lab Materials

| Category | Item | Qty / sample | Qty | Unit | Bulk qty | Price / bulk | Cost / Unit | Total cost / experiment |
|---|---|---|---|---|---|---|---|---|
| Consumable | Pipette tips | N/A | 562 | tips | 960 | $130 | $0.14 | $76.10 |
| Consumable | Tubes | N/A | 148 | tubes | 250 | $55 | $0.22 | $32.56 |
| Consumable | PCR plate | N/A | 1 | plate | 25 | $157 | $6.28 | $6.28 |
| Consumable | PCR plate film | N/A | 1 | film | 100 | $43 | $0.43 | $0.43 |
| Consumable | Gloves | N/A | 10 | gloves | 1000 | $268 | $0.27 | $2.68 |
| Reagent | Lysis buffer | 0.2 | 6.4 | mL | 500 | $111.84 | $0.22 | $1.43 |
| Reagent | Trizol | 0.8 | 25.6 | mL | 200 | $374 | $1.87 | $47.87 |
| Reagent | Chloroform | 0.12 | 3.84 | mL | 500 | 22.14 | $0.04 | $0.17 |
| Reagent | Glycogen (RNA grade) | 1 | 42 | uL | 200 | $81.25 | $0.41 | $17.06 |
| Reagent | 200-proof Ethanol | 2 | 84 | mL | 2000 | $109 | $0.05 | $4.58 |
| Reagent | Water (Mol Bio Grade) | N/A | 10 | mL | 1000 | $28.34 | $0.03 | $0.28 |
| Reagent (PCR) | PCR mastermix | 5 | 210 | uL | 500 | $204 | $0.41 | $85.68 |
| Reagent (PCR) | Primers | 1 | 42 | uL | 500 | $99 | $0.20 | $8.32 |
| Total | | | | | | | | $283.45 |

#### Sampling Materials

| Category | Item | Qty | Unit | Bulk qty | Price for bulk | Cost / Unit | Total cost |
|---|---|---|---|---|---|---|---|
| Sample Material | Filters max | 10 | Filter | 100 | $114 | $1.14 | $11.40 |
| Sample Material | Swabs | 15 | Swab | 100 | $190.72 | $1.91 | $28.61 |
| Sample Material | Cleaning wipes | 30 | wipes | 225 | $11.97 | $0.05 | $1.60 |
| Sample Material | Gloves | 40 | gloves | 100 | $18 | $0.18 | $7.20 |
| Sample Material | Baggies | 30 | bags | 300 | $7.12 | $0.02 | $0.71 |
| Sample Material | Secondary containers | 1 | Container | 6 | $52.19 | $8.70 | $8.70 |
| Total | | | | | | | $58.21 |

#### Code Availability

Code for processing PCR results will be made public on Github at: (url to be shared upon publication).

### Graphical TOC Entry

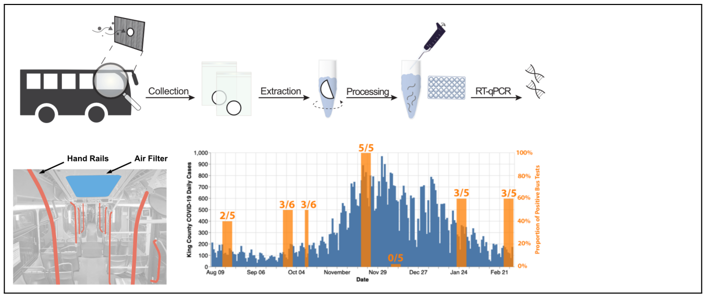


This journal requires a graphical entry for the Table of Contents. This should be laid out "print ready" so that the sizing of the text is correct.
Inside the `tocentry` environment, the font used is Helvetica 8 pt, as required by *Journal of the American Chemical Society*.
The surrounding frame is 9 cm by 3.5 cm, which is the maximum permitted for *Journal of the American Chemical Society* graphical table of content entries. The box will not resize if the content is too big: instead it will overflow the edge of the box.
This box and the associated title will always be printed on a separate page at the end of the document.
See here for more information: http://pubsapp.acs.org.offcampus.lib.washington.edu/paragonplus/submission/toc_abstract_graphics_guidelines.pdf?
